# Lessons learned from the resilience of Chinese hospitals to the COVID-19 pandemic: a scoping review

**DOI:** 10.1101/2021.03.15.21253509

**Authors:** Jack Stennett, Renyou Hou, Lola Traverson, Valéry Ridde, Kate Zinszer, Fanny Chabrol

**Affiliations:** CEPED, Institute for Research on Sustainable Development, IRD-Université de Paris, ERL INSERM SAGESUD, 45 Rue des Saints-Pères, 75006, Paris, France.; Centre de recherche en santé publique, Montréal, Canada / Université de Montréal, Montréal, Canada

## Abstract

As the SARS-CoV-2 pandemic has brought huge strain on hospitals worldwide, the resilience shown by China’s hospitals appears to have been a critical factor in their successful response to the pandemic. This paper aims to determine the key findings, recommendations and lessons learned in terms of hospital resilience during the pandemic, as well as the quality and limitations of research in this field at present.

We conducted a scoping review of evidence on the resilience of hospitals in China during the COVID-19 crisis in the first half of 2020. Two online databases (the CNKI and WHO databases) were used to identify papers meeting the eligibility criteria, from which we selected 59 publications (English: n= 26; Chinese: n= 33). After extracting the data, we present an information synthesis using a resilience framework.

We found that much research was rapidly produced in the first half of 2020, describing certain strategies used to improve hospital resilience, particularly in three key areas: human resources; management and communication; and security, hygiene and planning. Our search revealed that considerable attention was focused on interventions related to training, healthcare worker well-being, e-health/ telemedicine, and work organization, while other areas, such as hospital financing, information systems and healthcare infrastructure, were less well represented in the literature.

We identified a number of lessons learned regarding how China’s hospitals have maintained resilience when confronted with the SARS-CoV-2 pandemic. However, we also noted that the literature was dominated by descriptive case studies, often lacking consideration of methodological limitations, and that there was a lack of both highly-focused research on individual interventions and holistic research that attempted to unite the topics within a resilience framework. Research on Chinese hospitals would benefit from a greater range of analysis in order to draw more nuanced and contextualised lessons from the responses to the crisis.

## 1 Introduction

It is crucial to study the SARS-CoV-2 pandemic in terms of its impact on healthcare systems, health care facilities and health workers (WHO, 2020). Since its rapid introduction, the pandemic has posed serious problems for hospital resilience globally (Thomas et al. 2020), with effects ranging from over-occupation of ICU (intensive care unit) beds (Carenzo et al. 2020), overworking of medical staff while treating COVID-19 patients (Fan, 2020) to the inability to provide or underuse of other services (Søreide et al., 2020). In addition to the challenges of meeting increased capacity needs, healthcare systems and hospitals have had to prepare for and minimise the risk of nosocomial infection, which has often required major infrastructure and organizational changes (Xu et al. 2020).

The response of hospitals in China to the pandemic in early 2020, particularly the situation in Wuhan, has been well-publicised. As Wuhan was the source of the first large (documented) nosocomial outbreak, many feared that hospitals in the city and elsewhere in China would struggle to cope with the shock of the pandemic (Wang, Zhou, He et al. 2020). However, hospital strategies contributed to a concerted national effort, including a strict lockdown and forceful restrictions on movement and association, which allowed case numbers to become negligible by late March. The final COVID-19 patient in Wuhan was finally discharged on 5 June 2020 (Burki, 2020). Although China maintained certain restrictions throughout 2020 and experienced other minor outbreaks and suffered economic losses in the first half of the year, the country’s response has since been viewed as a comparative success story (Walker et al., 2020).

Resilience is defined as a system being able to adapt its functioning to absorb a shock and, if necessary, transform to recover from adverse events and has become an increasingly salient issue within international health and development literature (Turenne et al., 2019). However, resilience is used less frequently to address hospital and health system issues in the Chinese context. Zhong et al. (2014a) have performed a scoping review examining disaster health management, health infrastructure safety, disaster preparedness and medical response capability in China. They found that the topic was poorly covered in the Chinese context in both English- and Chinese-language literature.

Research by the same team (Zhong et al., 2014b) led to the development of a quantitative conceptual framework of hospital disaster resilience with a Chinese focus that highlights the role of hospital resilience in the first SARS pandemic in 2003. The study, based on questionnaires addressed to tertiary hospitals across Shandong province, identified four key factors that reflected the overall levels of disaster resilience (hospital safety, disaster management mechanisms, disaster resources and disaster medical care capability), and compared the extent to which hospitals in the region met these criteria. They found that there was major variability within the province under study (Shandong) based on the type and location of hospitals. While some elements of resilience were commonly achieved (93% of hospitals had infectious disease surveillance), others were only managed by certain hospitals (e.g. only 12% of hospitals were able to surge staff capacity). However, this literature must be reconsidered in the light of the recent SARS-CoV-2 outbreak, where the resilience of China’s hospital has been challenged by a major health crisis.

At the time of writing, many other countries are entering a second or third wave of the pandemic, new, potentially more contagious strains are rapidly emerging, and health systems are being increasingly and continually challenged. While the success of China’s strategy has been attributed to achievements of the government, public health officials, and the attitudes of the general public (Walker et al., 2020), the specific role of hospital resilience in this strategy is less documented. We, therefore, have conducted a scoping review to identify and synthesise the literature regarding the resilience of China’s hospitals in the context of the COVID-19 pandemic during the first wave and to draw lessons from these experiences to better inform and improve responses to the current pandemic and to future crises.

## 2 Methods

As part of a multidisciplinary team, and with the support of two librarians, we chose a scoping review to enable us to synthesise, with rigour and in a relatively short period of time, the state of knowledge on our research question and to identify and analyze gaps in the knowledge base (Munn et al., 2018). The scoping review was preferred to a full systematic review as our goal was to provide, from a very broad search, rapid information for public decision makers, stakeholders and researchers regarding insights into hospital resilience in China.

We conducted our review based on the PRISMA methodology (Moher et al., 2018), which is largely based on the methodological framework of Arksey and O’Malley (2005).

### 2.1 Protocol and Registration

In June 2020, we registered this study on protocols.io (https://www.protocols.io/edit/lessons-learned-from-the-resilience-of-chinese-pub-bjpckmiw).

### 2.2 Relevant literature identification

We conducted a systematic search using two different strategies to select appropriate academic literature from each context.

For the English-language literature, we have based our research on a collection of articles related to the COVID-19 pandemic published on the WHO website (https://search.bvsalud.org/global-literature-on-novel-coronavirus-2019-ncov/). These articles were collected from the following databases: Medline (Ovid and PubMed), PubMed Central, Embase, CAB Abstracts, Global Health, PsycInfo, Cochrane Library, Scopus, Academic Search Complete, Africa Wide Information, CINAHL, ProQuest Central, SciFinder, the Virtual Health Library, LitCovid, WHO COVID-19 website, CDC COVID-19 website, China CDC Weekly, Eurosurveillance, Homeland Security Digital Library, ClinicalTrials.gov, bioRxiv (preprints), medRxiv (preprints), chemRxiv (preprints), and SSRN (preprints).

The search terms included the following keywords, comprising the three concepts: 1) China; 2) Healthcare systems, Hospitals, and professionals; 3) Resilience. English-language search terms (Table 1) and methodology were checked by a professional librarian affiliated with the IRD. For the Chinese-language literature, we searched for relevant articles on the database CNKI using the following search terms (Table 2). The request in Chinese was designed using an iterative process in consultation with a Chinese-speaking librarian from BULAC (Bibliothèque Universitaire des Langues et Civilisations).

**Table 1:** Search Terms.

| Concept | China | Healthcare systems, Hospitals and Healthcare Professionals | Resilience |
| --- | --- | --- | --- |
| **Keywords** | China; Chinese; PRC; People's Republic of China; Wuhan; Hubei. | healthcare ; health care ; health system ; hospital ; health facilit ; health center ; medical center ; health service ; worker ; staff ; clinician ; personnel ; human resource ; professional ; volunteer ; physician ; nurse ; paramedic ; doctor_ ; doctors ; workforce ; trainee | resilienc ; shock ; crisis ; crise ; challenge ; emergenc ; disturbance ; capacit ; respons ; strength ; adapt ; strateg ; prepar ; readiness ; sustain ; effectiv ; stress ; impact ; effect ; surge ; extraordinary ; organization ; organisation ; optimi ; restructur ; communicat ; collaborat ; coordinat ; partner ; essential function ; basic function ; logistic ; service ; structural measure ; access ; resource ; equipment ; supply ; supplies ; medication ; drug ; policy ; policies ; governance ; leader ; manag ; financ ; funds ; funding ; training ; recruit ; innovat ; regulation ; triage ; evaluat ; support ; hopeless ; helpless ; efficien ; opportunit ; solution ; frontline ; engagement ; coping ; priorit |

**Table 2:**
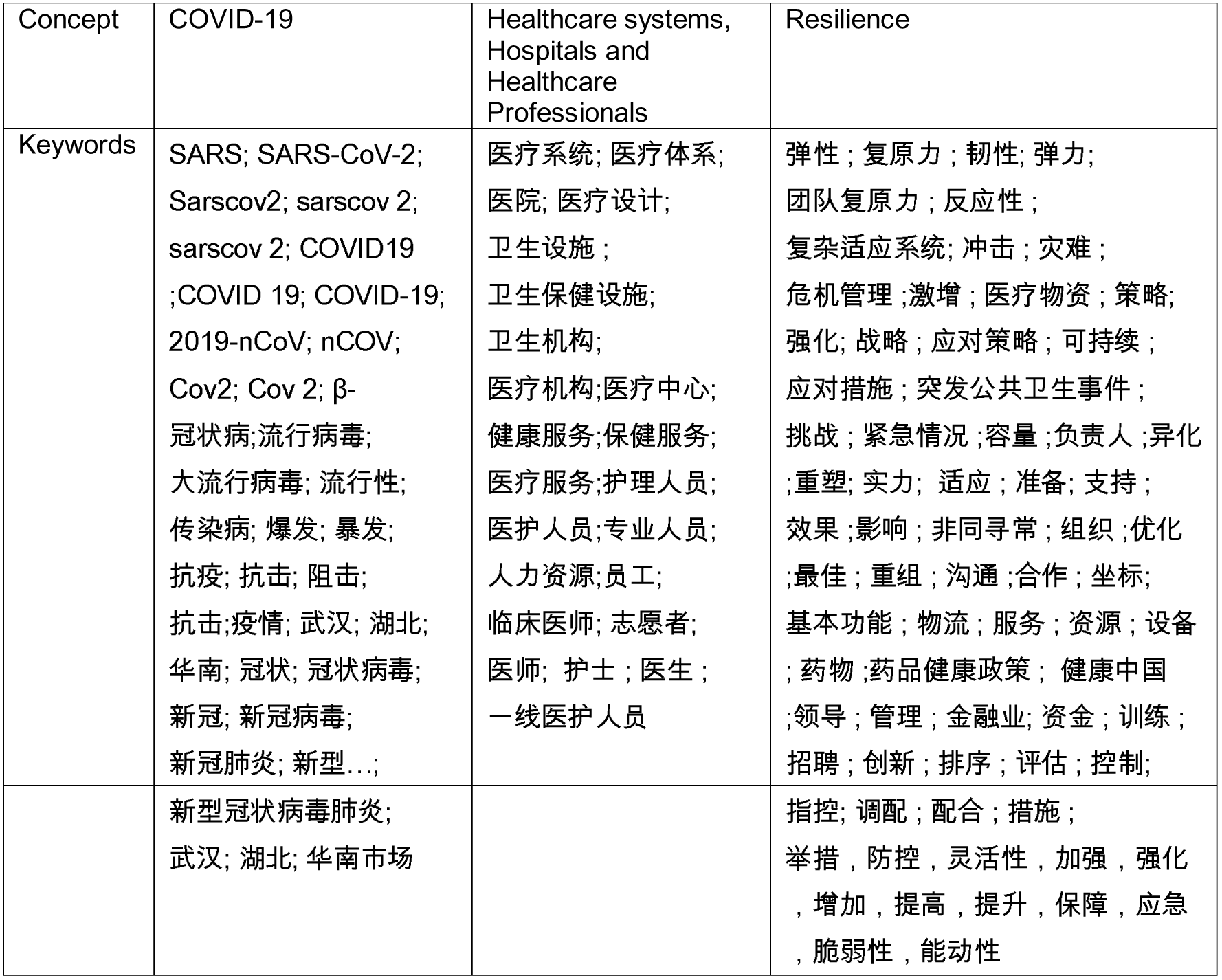
Search terms used in Chinese-language search.

In order to filter the results, we chose to limit the search on CNKI to five subcategories of article: those included in the ‘Science Citation Index’, the ‘Engineering Index’, the Beidahexin (Beijing University Core Journal Database), CSSCI (The Chinese Social Science Citation Index) and CSCD (Chinese Social Science Database).

The selection of sources of evidence was conducted following an extended iterative process, confirming the overlap of articles with searches on other platforms (e.g., Wanfang, Google Scholar, PubMed, CDC Website). We limited our searches to the WHO and CNKI databases after they were determined to be sufficiently comprehensive.

### 2.3 Data Extraction Process

The following information was extracted from each of the selected articles: title, authors, publication type, type of resilience, whether resilience was explicitly referred to, the hospital dimension, main objectives of the article, a slightly adapted Mixed-Methods Appraisal Tool (MMAT) evaluation, a simple representation of the results, limitations and main findings, recommendations by the authors, and some subjective notes by the reviewers. The MMAT tool was adapted to better capture single case studies (Atkins and Sampson, 2002).

### 2.4 Study Selection

Articles were included in the review if they: i) were published between December 2019 and July 2020; ii) were published in English or Chinese; iii) focused on the resilience of Chinese hospitals in the COVID-19 pandemic; iv) included empirical results; v) full articles were accessible, and vi) not considered grey literature (i.e. press articles, letters, editorials etc.).

Two reviewers (JS and RH) used the software Rayyan (https://rayyan.qcri.org) to select the papers using a two-stage review process. Articles were initially excluded based on titles and abstracts and then the full paper was evaluated. If an included article was identified as concentrating on public health systems, hospitals or healthcare professionals, it was classified as such, and only included in the study if it pertained to hospital resilience. ‘Public health system resilience’ pertained to elements that reflect broader choices made by the health system, such as media, supply chains and non-pharmaceutical interventions; ‘hospital resilience’ pertained to choices made by and within individual hospitals, and ‘healthcare professional resilience’ referred to the individual and group resilience of healthcare staff, such as psychological issues, physical injuries or exhaustion experienced by staff. There was a significant amount of overlap, therefore many studies were identified as belonging to more than one category (see table 5).

### 2.5 Critical appraisal of individual sources of evidence

Two authors (Anonymous) used MAXQDA® 2020 to code the data using a coding tree consisting of 7 larger categories: governance, human resources, professional values, finance, security, planning and management, communication, background (pre-existing policies), as well as two other coding categories to map methods (including methodological limitations) and the conceptual framework dimensions (see below). A separate category for professional opinions, recommendations and other cited articles was included to facilitate the synthesis.

The quality of studies was not evaluated although we did include information on the type of study design, data collection methods, potential limitations, a summary of the results, main findings and recommendations given within the articles (Table 5).

### 2.6 Data synthesis and the conceptual framework

Results from the Chinese- and English-articles were initially synthesised separately by RH and JS respectively, then the two syntheses were combined by all authors. We performed the synthesis according to the Ridde et al. (2021) definition of healthcare system resilience: “the capacities of dimensions/components of a health system faced with shocks, challenges/stress or destabilizing chronic tensions (unexpected or expected, sudden or insidious, internal or external to the system), to absorb, adapt and/or transform in order to maintain and/or improve access (for all) to comprehensive, relevant and quality health care and services without pushing patients into poverty”.

The synthesis of the articles was performed in terms of context (comprising background/context, as well as events and effects in the above framework), strategy and impact. Firstly, we explain the context in which a specific strategy is adopted, including the events in question and the effects of the pandemic experienced by the hospital in question. Subsequently, we describe a synthesis of the strategies, giving examples if necessary. Finally, we note the impacts of these strategies, which can theoretically be positive, negative or neutral. The causality attributed to certain interventions will be examined cautiously in the discussion section. The right side of this resilience framework (parts 3 and 4) is not used in the evidence synthesis, but will be examined briefly in the discussion section.

This framework also helps us to address the question of how hospitals anticipate or react to crises. The ‘effect-strategy-impact’ stage can illustrate different configurations: a) a reaction, when all three factors are present: an effect is felt, a strategy is adopted and this strategy has positive or negative impacts, b) anticipation, when strategies have a positive or negative impact, before a shock or independent of the effects of the shock, and c) inaction, when the effects are felt, but there are no strategies in place to react to the shock.

The framework identified 10 conceptual dimensions of health systems: governance, intervention level, workforce, culture and social values, finance, planning and supported guidance, systems specificities, health sector management, information systems, context and security, which we integrate into three larger categories with which to perform the synthesis: human resources, management and communication and the hygiene-security-planning nexus.

## 3 Results

As shown in Figure 1, we obtained 888 articles in Chinese and 5031 in English.

**Figure 1:**
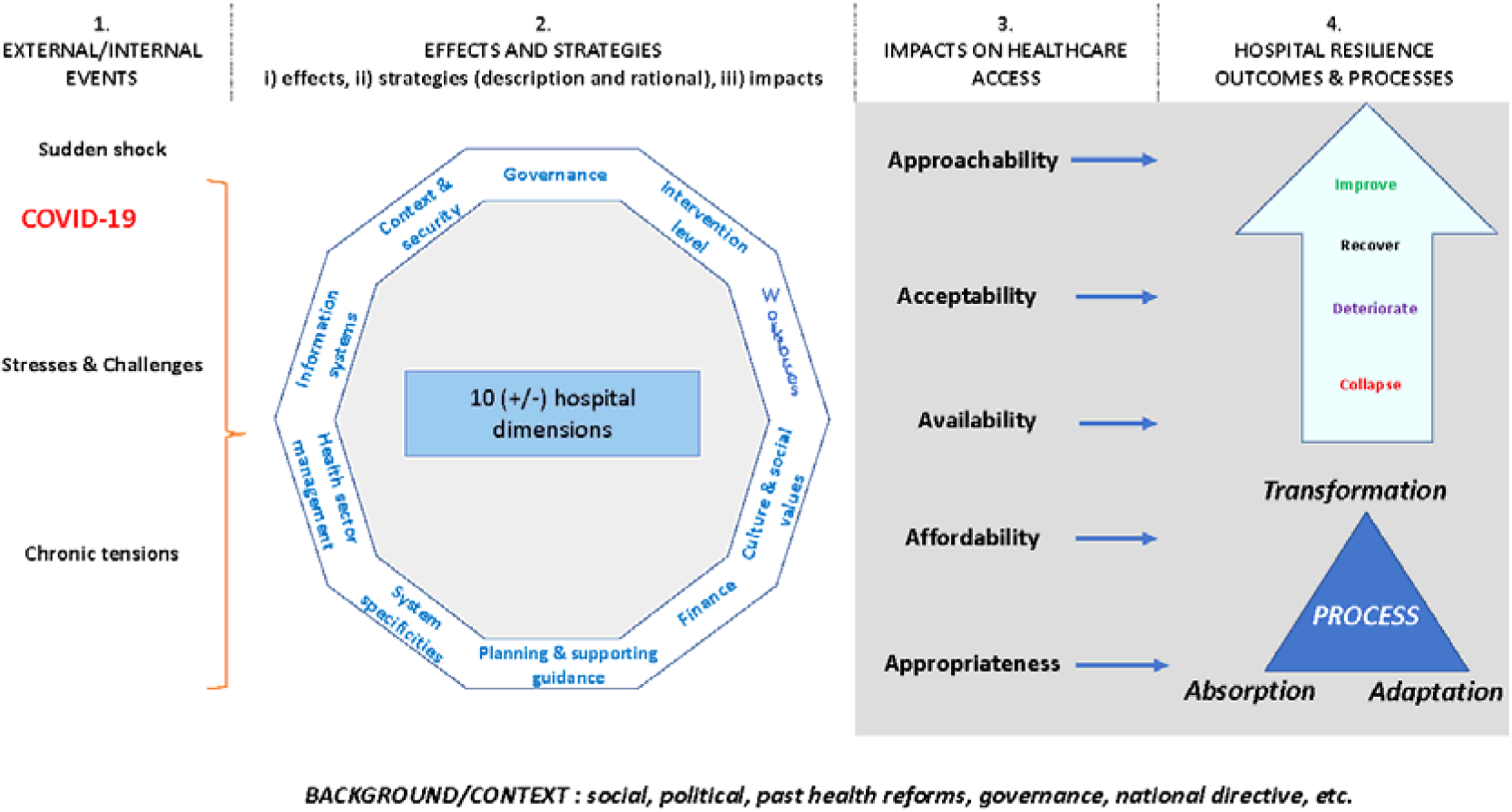
Conceptual Framework.

We identified 236 studies that met the criteria regarding resilience in general, of which 59 studies, 26 in English and 33 in Chinese, met the criteria for inclusion in the hospital-focused study; Figure 1 shows the process of study inclusion in this scoping review. We mapped the distribution of study design according to region, type of study, category of hospital and language (Table 3).

The geographical distribution of the papers is described in figure 3: the studies were based on research undertaken at a diverse array of hospitals and settings. Only 2 articles were based on national surveys, and therefore not focused on a single hospital. Understandably, the most represented geographical location was Wuhan, in Hubei province (24.1%), with Sichuan province in second place (13.8%), followed by Guangdong (12.1%) and Shanghai (8.6%). 84.5% of studies were focused on tertiary A hospitals, the highest-ranked large hospitals in the country; 13.8% of studies included various hospitals, including secondary hospitals, while only 1 study (1.7%) targeted primary healthcare providers.

**Figure 2:**
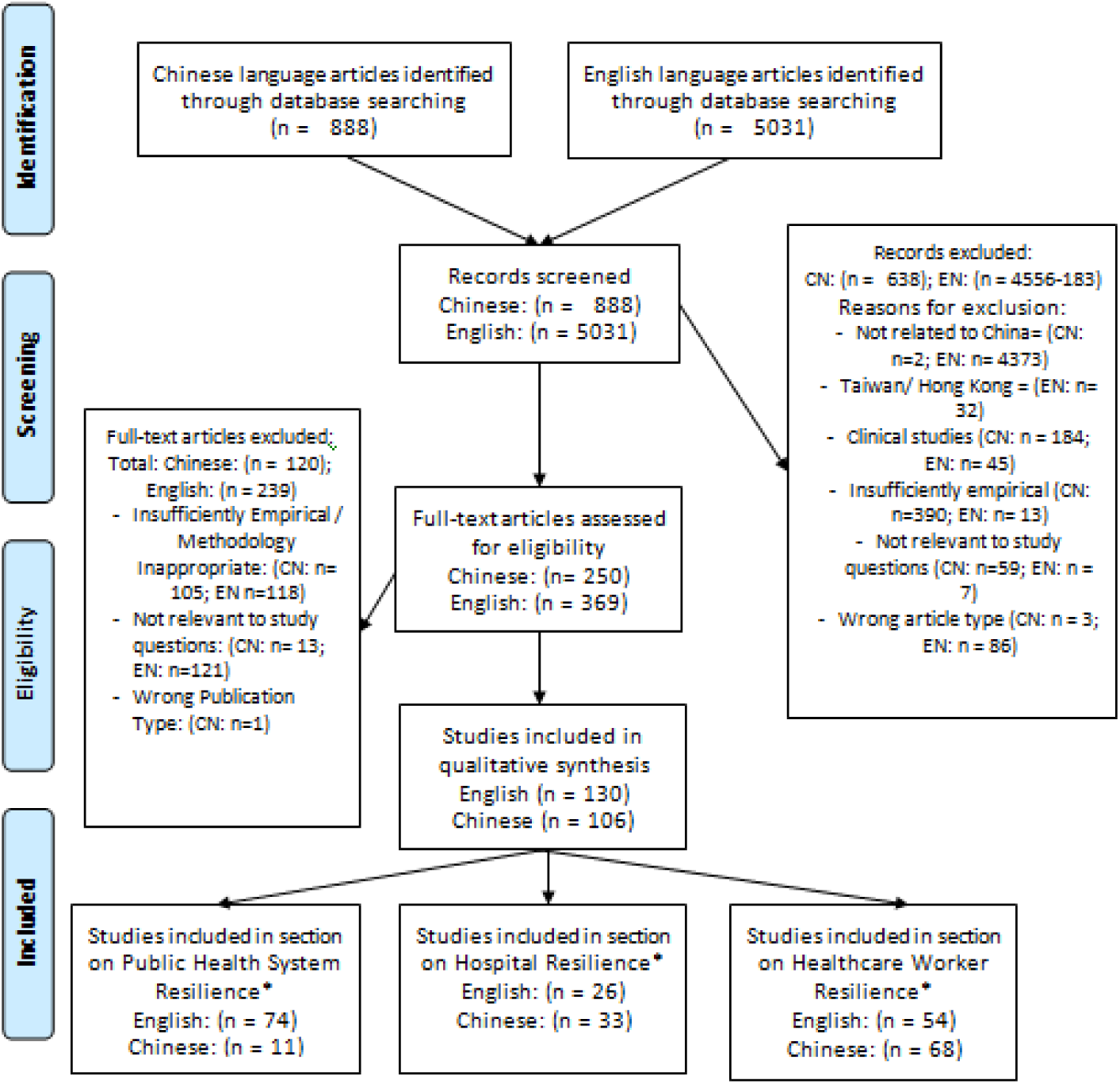
Flow Diagram.

**Figure 3:**
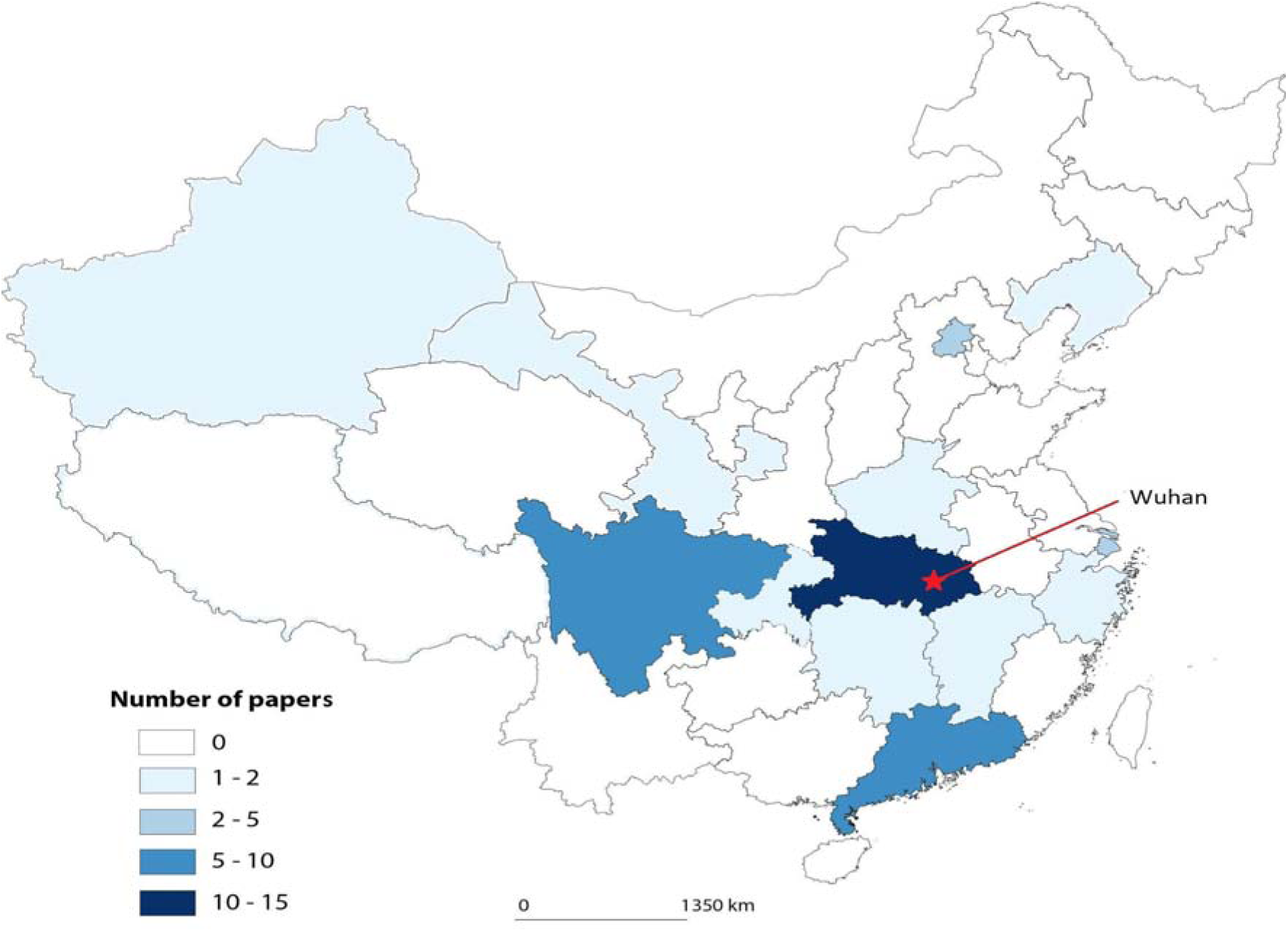
Geographical distribution of included studies at the Provincial level in China.

Our analysis revealed that 91.5% (n = 54) of the articles were identified as peer-reviewed articles, with a single review article, a commentary article, a short report, and two articles identified as “Peer-Reviewed Article/ Quality Improvement Study” and “Peer-Reviewed Article and Review” respectively. In terms of methodology, the studies were dominated by single case studies using mixed methods (50.8%; n = 30) and descriptive quantitative studies (37.3%; n = 22). There were only 4 qualitative studies (6.7%), 2 studies using other mixed methods (3.4%) and 1 randomised study (1.7%). The dimensions of hospital resilience most commonly referred to were health sector management (n= 44), context and security (n = 47), intervention level (n = 8), planning and support (n = 29), system specifics (n = 8), information systems (n = 8), workforce (n = 31), and cultural and social values (n =4). Other dimensions such as governance and finance, were not covered in the selected articles.

In terms of MMAT criteria, 84.7% (n = 50) of articles contained clear questions and objectives, and addressed them appropriately. The characteristics of quantitative studies were mixed in terms of adherence to the MMAT criteria: the sampling strategies were often not made explicit (n = 4) and many studies used some form of convenience sampling (n = 5) due to accessibility and need for timeliness given the crisis context. It was often unclear whether the study was representative of the population (n = 8) and in some articles (n = 6) there appeared to be some overrepresentation of certain groups within the population (women and nurses in particular).

All quantitative articles were deemed to have used appropriate measurement tools, and only 9.0% (n = 2) of quantitative articles did not specifically mention response rates. In the mixed methods articles, 6.7% of articles (n = 2) were considered to have used an unclear methodology and 16.7% (n = 5) were identified as not integrating qualitative and quantitative data in a relevant manner. 35.6% of articles explicitly considered limitations of the study methodology.

Studies published in Chinese were more likely to be case studies (n = 21; 61.7%) compared to studies published in English (n = 7; 26.9%). Chinese-language studies were also less likely to consider limitations of the methods used, with 70.6% (n = 24) not mentioning any limitations, as opposed to 23.1% (n = 6) of the English-language papers. Our MMAT evaluation determined that the papers published in English conformed more closely to the norms established in the MMAT framework than those in Chinese.

We identified 10 different categories in which strategies to address the pandemic were employed, and 29 specific strategies recommended by the articles (Table 4), with a description of the strategies involved and in which studies these strategies were mentioned. Table 4 illustrates that a number of different strategies were identified in the scoping review procedure, and the results of these are explored in the following synthesis.

### 3.1 Human Resources

#### 3.1.1 Reinforcements

##### Context

After the initial outbreak of COVID-19 in Wuhan, the Chinese authorities made the decision to send personnel reinforcements from all over the country to Hubei province to fight the epidemic. Wuhan is medically well-equipped (9.25 hospital beds per 1,000 inhabitants in 2018) and has 110,000 health professionals, including 40,000 medical practitioners and 54,434 nurses (Wang, 2020). At the end of January 2020, reinforcements were nevertheless sent from all over the country: 42,000 new health workers arrived in Hubei province, including 35,000 in the city of Wuhan alone. They remained in the area for between 18 and 50 days (Guiheux et al., 2020). The implementation of a rapid response mechanism to the pandemic thus requires the rapid integration of external reinforcements sent to Hubei and their adaptation to the local hospital environment and management style (Wang, Yang et al. 2020). Several articles reported on the strategies implemented by hospitals to facilitate the integration of reinforcements and improve work efficiency (Zhao, Liu, Li et al. 2020, Han, Deng et al. 2020).

- **Strategy 1: Standardization of nursing procedures.** In one hospital in Wuhan, the 27 nurses in a department dedicated to COVID-19 patients had all come from different departments (infectiology, cardiology, internal medicine, etc.) in six different hospitals from within Sichuan province, demonstrating a wide diversity of backgrounds. As nurses did not have the same working experiences, skills, or working habits, in order to facilitate collaboration and improve work efficiency, the management introduced a standardised work system regarding nursing procedures for the workflow, work content, and responsibilities of each staff member (Feng, Wang, Su et al. 2020).
- **Strategy 2: Creation of back-up teams.** This involved staff members and external reinforcements in preparation of increased staff demand in the COVID-19 infection wards and/or to compensate for a loss of staff due to infection. For example, Jinyintan Hospital in Wuhan deliberately split nursing teams into teams comprising both: 1) back-up (non-local) nurses and local nurses; 2) experienced nurses and newly graduated nurses; 3) intensive care unit nurses and non-intensive care unit nurses, in order to share experience, skills and awareness of procedure (Guo et al, 2020).
- **Strategy 3: Delineate the responsibilities of each staff member.** Many hospitals instituted measures, such as checklist interventions, training and management strategies, to ensure that the roles and responsibilities of all staff, especially transdisciplinary nurses and new staff, were clearly defined and that staff were aware of any and all changes. These new responsibilities included recording ECG results, organizing ward supplies, and observation of critical patients (Liu et al., 2020). Training interventions concerned both protective measures and the operation of medical equipment. As the back-up nurses were not familiar with emergency equipment and instruments, hospitals set up a series of training programmes to help the nurses understand the operation of various pieces of equipment (Feng, Wang, Su et al. 2020).

##### Impacts

The articles reported many positive outcomes as a result of these interventions, including how the efforts helped facilitate the integration of the reinforcement into the service and deliver quality care with efficiency, while maintaining the mental and psychological health of reinforcement staff (Guo et al., 2020, Wang, Yang et al., 2020). For example, Feng, Wang, Su et al. (2020) described how, between 27 January and 15 March 2020, as a result of the close collaboration between local nurses and back-up nurses, 86.6% of the COVID-19 patients admitted to the hospital were discharged (by the time of writing), and the hospital had only suffered one fatality. Also, the care and psychological support offered to both back-up and local nurses was timely with none of their nurses having reported any severe psychological problems. Additionally, as Liu, Cheng et al. (2020) note, reinforcement staff had negligible levels of infection likely due to strict adherence to procedures, ample access to PPE equipment, and special accommodations away from their families and other potential sources of infection.

#### 3.1.2 E-health, telemedicine and use of technology

##### Context

Due to the contagiousness of SARS-CoV-2, new ways of caring for patients were implemented to reduce the risk of contamination. Family visits were restricted and the loneliness of inpatients became a major challenge, requiring hospital staff to pay more attention to the mental health of patients. In order to prevent and control the spread of the virus and to avoid cross-contamination, some departments closed their ambulance services and stopped receiving patients. A large number of patients receiving radiotherapy, chemotherapy, or dialysis could not be treated in a timely manner (Liang, Deng et al. 2020). In addition, lockdown and the accompanying traffic control measures made it difficult for non-COVID-19 patients to travel and receive treatment (Luo, Chen et al. 2020).

In order to continue providing healthcare to the community and fulfil their obligations to patients, hospitals had to use other methods to provide care to patients with or without COVID-19. For this reason, a common element examined in the chosen studies was the use of telemedicine interventions. Telemedicine interventions serve the role of: 1) Allowing patients to receive medical appointments, services and treatment without having to visit a hospital, 2) Preparing and screening patients before they arrive at the hospital, to facilitate their entry into the hospital and avoid contamination, 3) Monitoring COVID-19 patients in home quarantine, and, 4) Providing more efficient use of human resources. Additionally, mobile and digital technology was used by hospitals across China in a range of other ways to increase efficiency and reduce person-to-person contacts (Chen et al., 2020).

- **Strategy 1: Using online services for psychological issues in the population**. A hospital in Chengdu implemented a multi-tiered intervention program, with online media, free hotline consultation, and online video interventions provided to citizens with psychological problems, with crisis intervention provided on-site (He et al., 2020,).
- **Strategy 2: Developing online screening mechanism for potential patients**. A range of strategies were suggested to provide web-based consultations, appointments, prescription services and drug delivery, and other services, as a complement to the physical hospital services. For example, in a qualitative study of patients’ experiences with online services offered to non COVID-19 patients, one patient reported: “The use of mobile apps in this pandemic is very important. You can pay, register, and view results on your mobile phone. You don’t need to queue up at the outpatient clinic, and you can chat with a doctor online after you get home, so it’s far more secure.” (Li, Huang, Zhang et al. 2020)
- **Strategy 3: Using online platforms to monitor COVID-19 patients.** A number of articles described a process of offering ‘e-counselling’ support to patients who were struggling with the physical and psychological effects of the disease (Hao et al., 2020). In addition to providing a greater monitoring and awareness of the individual patients, this intervention also allowed the staff to collect data to use in the improvement of in-hospital treatment (Yang et al., 2020, Shao et al., 2020).
- **Strategy 4: Developing and using onsite IT services and infrastructure.** The use of non-medical technology to improve hospital services, such as using app-based QR codes (unique Quick-Response codes, a machine-readable optical label, similar to a barcode, often produced by a smartphone) to share information and using robots for certain tasks to avoid person-to-person contact, was expanded during the pandemic (Chen et al., 2020).

##### Impacts

The effectiveness of telemedicine interventions was reported in many papers and largely viewed as an effective substitute or complement to onsite healthcare (Shao et al. 2020). They were also cited as being popular among the majority of users in a single study (Li, Chan et al., 2020). The introduction of teleconsultations reduced the difficulties of patients with chronic illnesses regarding the management and purchase of medicines (Luo, Chen et al. 2020) and the time required for consultation was also further shortened using teleconsultation (Li, Chan et al. 2020). Furthermore, the implementation of e-health interventions allowed staff to address the needs of a higher number of patients, reduced hospital load, while also helping to spread understanding regarding the virus risks and public health knowledge (He et al., 2020, Huang et al., 2020).

#### 3.1.3 Organisation of Work and Healthcare Worker Well-Being

##### Context

At the beginning of the epidemic, medical personnel suffered from panic and fear due to insufficient knowledge of the epidemiological characteristics of the virus and the need for protection, as well as experiencing a temporary shortage of medical supplies (Zhang, Shi et al. 2020). The problems of work overload were also highlighted in many papers, particularly the problems associated with new, unfamiliar tasks for which nurses had not been specially trained (Wu et al., 2020), high work intensity (Lai et al., 2020), disrupted circadian rhythms, as well as restrictions and challenges of protective clothing.

These factors intensified workload pressures and led to anxiety, insomnia, depression, pain, symptoms of PTSD, and grief. Furthermore, a higher workload led to worse hygiene behaviour, such as reduced adherence to hand-washing guidelines (Li and Chan et al., 2020). As one article explains: “high risk of professional exposure, high workload, transmission of patient anxiety, helplessness towards ineffective treatment of some severely ill patients, etc. can lead to high levels of psychological pressure, low confidence in one’s own work and depression among nurses, which affects the quality of work and the physical and mental health of nurses” (Feng, Wang, Su et al. 2020).

- **Strategy 1: Promoting readjustment of healthcare staff schedules**. For healthcare staff who had direct contact with COVID-19 patients, a 4- to 6-hour schedule was put into place by a number of hospitals. The protection protocols in Chinese hospitals were extremely strict, especially for health care workers who were working directly with COVID-19 patients. Once PPE was applied, it was required that the wearer avoid all potential contamination risks, including physiological needs: eating, drinking, and using the toilet (Zhao, Li et al. 2020). The hospital thus reorganised the timetable of the staff, accounting for the physical condition of the caregivers, the effective usage of single-use protective material, and the level of needs of the patients. For example, in Jinyintan hospital (Wuhan, Hubei), three systems were successively implemented as early as December 2019: work hours were initially limited to 6 hours per day, then 4 hours per day and ultimately 5 hours per day (Guo et al, 2020). After having surveyed the staff’s perceptions of the three timetabling systems, the hospital adopted the 5-hour per day system. In another hospital in Wuhan, the hospital put into place 6 shifts per day with a 4-hour rotation to allow nurses the time to take care of their physiological needs (Han, Deng et al. 2020).
- **Strategy 2: Increasing flexibility of working hours according to the number and condition of inpatients.** During the high peak period of patient admissions, the number of staff was increased to provide an appropriate nurse-to-patient ratio, which is essential to ensure that patients receive appropriate care and that the workload of caregivers or staff remains reasonable. For example, in a hospital in Wuhan, each nurse was responsible for 6 to 8 patients (Han, Deng et al. 2020).The hospitals also used backup teams, while using shorter shifts and appropriate working hours to reduce risks associated with workload, including lowered quality of work, medical errors, and increased rates of nosocomial transmission (Guo et al, 2020, Li and Chan et al., 2020, Hao, Zhou et al., 2020).
- **Strategy 3: Providing material and psychological support to the staff**. As well as ensuring provision of essential supplies, several hospitals provided nutritional meals in order to support staff and improve their health and immunity. Many strategies were employed to provide psychological support, either through colleagues, health professionals, or specialised psychologists. For example, a maternity ward in Tongji Hospital (Wuhan) established a WeChat group, “to promote scientific articles on mental health, to understand in a timely manner, problems in the life and work of the medical staff and subsequently to provide help and support. (…) A psychological consultation platform was also set up to provide medical staff a channel to vent their negative emotions and to offer psychological interventions when needed” (Yang, Zhang et al. 2020).

##### Impact

These strategies enabled better working conditions for healthcare workers and better quality of care for patients. A number of articles (Lin et al., 2020; Hao, Zhou et al., 2020) highlighted that use of shorter shifts and appropriate working hours could be effective strategies to deal with mental health needs, work quality, and hygiene requirements. Furthermore, a flexible work schedule also meant that staff members were less affected by fatigue and stress. For example, in Tongji hospital in Wuhan, 96.9% of the staff members were satisfied with the schedule of 5 hours per day, compared to 58.5% satisfaction with the previous schedule of 6 hours per day (Guo et al, 2020).

#### 3.1.4 Management and communication

##### 3.1.4.1 Emergency team and nursing management

###### Context

After the COVID-19 outbreak in Wuhan, China, human resources were rapidly reorganised within hospitals, between hospitals, and throughout the country.

Transdisciplinary nurses without specific expertise in infectious diseases were brought in to support COVID-19 wards (Wu et al., 2020), back-up teams were introduced, and frontline nurses had major responsibility changes. However, many issues arose from this reorganization, such as nurses lacking understanding about their specific responsibilities (Ding et al., 2020). Several strategies in management were employed to increase the effectiveness of medical staff under these new circumstances:

- **Strategy 1: Creation of new teams**. The majority of HR interventions mentioned in the articles took place shortly after the pandemic was announced in January. These involved the creation of new teams, such as the ‘nursing technical support team’ comprised largely of head nurses from different departments (Lin et al., 2020) and the ‘emergency management and sensing control team’, with a focus on procuring new information about the virus, and establishing an emergency management plan (Zhu et al., 2020).
- **Strategy 2: Implementation of Plan-Do-Check-Act (PDCA) cycle, a management tool.** This consisted of a repeated four-stage model for continuous improvement in quality management (Liu et al., 2020). In terms of HR management, this included 3 relevant components: defining the staff’s role and responsibilities, establishing a clear staffing structure and changing the shift handover modes, and testing and verification of procedures, such as evaluating nursing staff with questionnaires.
- **Strategy 3: Implementation of regular training for healthcare workers.** Given the rapidity of SARS-CoV-2 spread, healthcare staff required rapid training to properly apply the protection protocols, and also needed continual information regarding the evolution of knowledge about the virus. In addition, reinforcements who were unfamiliar with the workplace also needed to familiarise themselves with their new colleagues and the work environment. Many hospitals in our study implemented a dual training system: online training and face-to-face training on topics such as: “COVID-19 hospital infection prevention and control, hospital air purification management specifications, medical institution disinfection technical specifications, and personal protection requirements for disinfection and isolation” (Wu et al. 2020) was undertaken in order to improve care and reduce the risks of contamination between colleagues. The training content included the following elements: the characteristics of the service and the environment, spatial planning and reorganization, disinfection measures and knowledge of protection protocols, work procedures and the use of medical equipment. As well as training, WeChat groups were established to communicate up-to-date information on the progression of the pandemic and knowledge of treatment options (Yan et al. 2020).

###### Impacts

Several articles quantitatively measured the effectiveness of different aspects of management interventions, finding that they succeeded in making staff aware of their roles and responsibilities, as well as clarifying the staffing structure and handover procedures (Liu et al., 2020, Zhao, Liu, Li et al. 2020). Through training interventions and communication facilitated by the WeChat groups, front-line caregivers developed a better knowledge of the virus, which helped to alleviate their anxiety and fear (Wang, Yang et al., 2020, Dong, Hu, Zhou et al., 2020), and they were better able to apply the protection protocols (Zhao, Guo, Sun et al., 2020). For example, a quantitative study focusing solely on a training intervention on COVID-19 knowledge and training techniques (Yan et al., 2020), found strong positive effects on employee knowledge, concluding that interactive simulation training is complementary to didactic teaching. In a hospital in Beijing, 7 days after the implementation of the standardised training program, a knowledge score related to the prevention and control of nosocomial infection rose from 69.0% to 88.5% after the training for 1,125 medical staff. The correct answers by supervisors rose from 82.7% to 92.3%, while the proportion of wearing surgical masks increased from 85.4% to 92.9%, and the proportion of adherence to hand hygiene protocols increased from 92.3% to 96.3%. (Wu, Guo et al. 2020). In Liu et al. (2020) the PDCA team identified the problem of poorly defined responsibilities, noting that, while 12.1% of staff initially lacked awareness of their responsibilities, this was reduced to 0% following a training intervention.

##### 3.1.4.2 Communication and information

###### Context

During the early stages of the outbreak, as knowledge of the virus progressed and the number of patients in the hospital increased daily. Hospitals were required to react immediately to the situation and readjust strategies accordingly, whether in terms of protection protocol, patient care, or work organization. The situation was more complex in hospitals with external reinforcement from others provinces because, according to one article, “each medical team has its own process and philosophy of care, the only way to provide quality care to patients is to coordinate and standardize and homogenize care” (Dong, Hu, Zhou et al. 2020). Quickly and accurately conveying expert information and the response plan to staff at all levels became a serious challenge for various medical institutions. In this situation, fluid communication between the different parties involved (government, hospital, carers, patients and families, etc.) was essential.

- **Strategy 1: Implementing regular meetings between the different team members for daily briefings.** For example, in Tongji Hospital in Wuhan where national medical aid teams served in a “whole-system-takeover model”, nursing department staffers worked in partnership to establish a range of measures including smoothing communication channels through daily meetings. According to one author: “In the early stages, we held daily nursing council meetings to shorten the adjustment period and standardise the work in order to shift from a ‘wartime state’ to a daily routine.” (Xu, Liu et al. 2020)
- **Strategy 2: Promoting the use of new information and communication technologies to aid communication between colleagues.** Use of communication platforms, usually WeChat groups, and occasionally telephone exchanges, was identified in a number of articles. In Tongji Hospital, in order to provide an effective communication and information mechanism, a WeChat group with all the nursing staff was set up to enable communication at any time. In addition, the hospital set up a daily nursing information system: the progress of nursing work as well as problems encountered in the quality control of care were analysed and then sent to everyone in image/text form (Xu, Liu et al., 2020).
- **Strategy 3: Promoting the use of visual materials to better convey information to healthcare staff**. This involved the use of physical signs such as multicoloured arrows indicating the different hospital zones and posters of protection protocols displayed in different zones (Wang, Yang et al., 2020)

###### Impact

Some evidence is provided that the communication measures mentioned above were effective in improving the psychological health and efficiency of healthcare workers. In the People’s Hospital of Wuhan University, one week after a ‘visual management’ communications intervention was implemented, the time taken to obtain materials was shortened, the satisfaction rate of medical staff improved and nursing quality increased (Wang, Yang et al. 2020). Moreover, updating regularly of knowledge and informing the caregivers promptly about key information, identified as: ‘what we know’, ‘what we don’t know yet’, ‘what we have [in the hospital]’, ‘what we don’t have [in the hospital]’ and ‘what we are doing’”, was seen as particularly effective, both in relieving the anxiety of healthcare workers and improving the effectiveness of protection measures (Wu, Guo et al. 2020, Xia, Li et al. 2020, Dong, Hu, Zhou et al. 2020).

#### 3.1.5 Security, Hygiene and Planning

As well as analyzing the number of infected staff, a large number of articles in the scoping review examined the reasons for infection of healthcare workers and presented hospital strategies to reduce the risk of nosocomial infections.

##### 3.1.5.1 Protection Protocols (change and application of protocols)

###### Context

The issue of contamination risk is one of the most frequently discussed topics in the articles and is related to many dimensions of hospital resilience, such as human resources, management, communications and information. The risk of nosocomial infection was extremely high in Wuhan, especially in the early phase of the outbreak. One study found that 84.5% of 1,688 infected health personnel believed that their infection had been acquired in the hospital wards (Gao, Sanna et al., 2020). In order to reduce the contamination risk as much as possible, Chinese hospitals implemented strict protocols for hospital admissions, discharge procedures, personal protective equipment, and in the application of social distancing rules.

- **Strategy 1: Strict management of hospital space**. In Lu, Hao et al (2020), access to the hospital was closely controlled in terms of body temperature and mask wearing. Patients with a temperature over 37.5°C and/or showing respiratory symptoms were redirected to the ambulatory service dedicated to fever patients or to the Emergency Department.
- **Strategy 2: Focusing on environmental contamination with routine disinfection**. In the COVID-19 unit, strict measures were applied regarding the disinfection of medical instruments (such as stethoscopes, thermometers, etc.). This was described in detail in one article, which explained how floors, tables, chairs, diagnostic and treatment beds were wiped and disinfected regularly with 1000 mg/L of chlorine disinfectant, and that this behaviour was regularly monitored (Liang, Deng et al. 2020).
- **Strategy 3: Encouraging healthcare workers to apply personal equipment protocols appropriately, according to their role and their level of contact with COVID-19 patients**. To help caregivers in properly applying the protocols, hospitals proposed regular training for staff and the establishment of a 24-hour supervisor position to verify the appropriate application of protocols when entering and leaving the buffer zone. Hospitals often introduced comprehensive management plans involving screening, personnel management, disinfection and hygiene procedures, training and supervision of employees, and PPE supply chains.
- **Strategy 4: Restricting family visits to avoid patient-family contact**. While family visits increased the risk for nosocomial transmission, many hospitals implemented a video visit system to facilitate exchanges between patients and their families, for example: “For people who come to the hospital to visit patients, the warden enables the video visit with an iPad connected to the nurses’ iPad at the patient’s bedside, which enables exchange with the visitors.” (Zheng, Liu et al. 2020)

###### Impact

Only a few articles evaluated the impact of these strategies on infection rates with most concluding that no medical staff member was contaminated by SARS-CoV-2 during this period. With regards to PPE use, a regression analysis in self-reported compliance with security protocols (Li and Chan et al., 2020) found two seemingly contradictory findings: while staff in high-risk departments have higher rates of compliance with security protocols, further contact with at-risk patients had a negative effect on compliance. The research did not capture information to determine whether this was due to resource shortages, human deficiency, high workload or other factors, but reducing workload through reinforcements, ensuring resource supply and increased training is recommended as an intervention. In terms of environmental contamination, an article (Jin et al., 2020) found the highest rates of environmental contamination in the isolation ward for pregnant women, even when compared to the fever clinic. It was hypothesised that this was due to the differences in ventilation and the number of visitors. Additionally, it found that hand sanitizer dispensers and used gloves were greater sources of risk of contamination than were eye protection or face shields.

##### 3.1.5.2 PPE (Personal Protective Equipment)

###### Context

In the response to the COVID-19 outbreak, the supply of personal PPE was a major challenge globally. This problem was also present in China, where several regions suffered from a shortage of PPE and disinfection products (Xia, Li et al. 2020). Hospitals were required to adjust the variety and quantity of protective materials in a timely manner to find an ideal balance between the level of equipment consumption and storage capacity, which was essential to ensure continuity of care.

**Strategy 1: Implementing an inventory register for important materials while standardizing the process of managing and using these materials.** In many hospitals, a physical security team leader was put in charge of recording the real-time use of equipment and strictly controlling the receipt and distribution of materials (Yang, Zhang et al. 2020).
- **Strategy 2: Avoiding over-consumption and waste of materials.** The presence of the hygiene team and the supervisor in the application of caregiver protocols was to make sure that caregivers wore PPE correctly and also that they avoided overuse of the PPE (Medical Working Group, 2020). In another hospital, when restocking materials, it was not permitted to mix materials with different expiry dates to ensure that different materials were used in order of expiry dates, from oldest to newest, to avoid waste due to expired materials (Wang, Yang et al. 2020).
- **Strategy 3: Decreasing the level of protection of certain non-COVID-19 services in view of the shortage of medical resources.** Some studies examined decreased protection measures to identify at the minimal level of protection needed. For example, a study in the Huaxi Hospital of Sichuan University suggested that the staff in non-COVID- 19 intensive care units did not need to wear full body protective overalls, thus saving on PPE (Wang, He et al. 2020).

###### Impacts

Over-consumption of PPE was a common problem in hospitals, particularly in the early and mid-stages of the outbreak. However, according to surveys from a hospital in Shenyang, the strategies mentioned above contributed to the optimization of PPE and disinfection supplies, allocating based on needs and stock, while ensuring that front-line personnel were well-protected (Zhang and Liu, 2020).The Huaxi Hospital of Sichuan University applied these protective protocols to its paediatric intensive care unit, finding that healthcare professionals (91 people) and household members (5 people) in contact with 91 COVID-19-positive patients wore only masks, and did not wear the full protective suit required by some institutions, yet there were no infections in the ward (Wang, He et al. 2020), suggesting that lighter PPE could be sufficiently effective, as well as saving material.

##### 3.1.5.3 Reorganisation of services

###### Context

During the outbreak, hospitals had to reorganise their services in order to both increase capacity and reduce the risk of contamination. The changes in infrastructure, hospital procedure and protocols in Chinese hospitals involved major changes. For example, a hospital in Wuhan revised 32 items on its regular hospital procedures in order to transform a general hospital into a designated COVID-19 treatment hospital (Lin et al., 2020), and a number of case studies were written on particular changes in procedure and ward renovation (Xu et al., 2020).

- **Strategy 1: Transformation of non-COVID-19 hospital areas into specialised COVID-19 wards.** Many hospitals lacked a negative pressure chamber; therefore the non-COVID-19 hospital area was required to be transformed into a specialised COVID-19 ward to accommodate the growing number of COVID-19 patients (Medical Working Group, 2020).
**Strategy 2: Reorganisation of space.** In designated COVID-19 hospitals, necessary infrastructure changes were implemented, which included setting up fever tents, ward renovation, unidirectional channels for patients, and converting sections of the hospital for COVID patients. These were done to minimise contact between infected and uninfected individuals, reduce patient flow throughout the hospital, and to maximise the shared space of COVID patients. Another example is the creation of the “three zones and two passages” system (Chinese:三区两通道) that included a contaminated zone, potentially contaminated zone, and a clean zone, as well as two separate passages for medical staff and patients.
- **Strategy 3: Reorganization of inpatient rooms.** Certain hospitals decided to convert double rooms into single rooms, whereas for hospitals that were forced to put several patients in the same room, a distance of more than one metre between beds was maintained.

###### Impacts

The impacts of these strategies were not examined in detail in the included studies. Many articles, such as Gao et al., (2020) claimed that their management strategy contributed to effective prevention of virus spread in the endoscopy centre in Sichuan; however, none of the articles claimed to provide strong evidence of the effectiveness of a given intervention.

## 4 Discussion

### 4.1 Summary of Evidence

In this scoping review, we identified 59 studies that address resilience in hospital settings across China in the context of the initial SARS-CoV-19 outbreak in the first half of 2020. Our findings indicate a wealth of research describing certain strategies used to improve hospital resilience, particularly those concerning the following dimensions: human resources; management and communication; security, hygiene and planning. We found that a great deal of attention was focused on training, healthcare worker well-being interventions, e-health and other technology related interventions, and work organization interventions, while training and management interventions were also subject to more rigorous quantitative analysis. Some themes, such as information systems and reinforcements, were mentioned in a small number of studies and lacked rigorous analysis while others, such as hospital financing and the development of new healthcare infrastructure were neglected in the literature, despite being mentioned explicitly in Chinese official policy papers (State Council Information Office of the People’s Republic of China, 2020). Most importantly, our findings also represented a paucity of rigorous research focusing on the effectiveness of interventions and a lack of research attempting to provide a comprehensive portrait, to unify these different elements within a resilience framework.

We have presented the results of the scoping review following the theoretical framework of “Effects – Strategies – Impacts” in order to study the resilience of Chinese hospitals. Only a handful of studies examined cases of inaction, for example Gao, Sanna et al. (2020) analyzed the reasons for personnel infections in the early stages of the outbreak. Most studies referred to actions taken in anticipation of major outbreaks in provinces with only limited spread. Some studies, especially those carried out within Wuhan, described a strong reaction to a serious ongoing outbreak. The majority of the included studies provided details on the effects and strategies with an appropriate methodology, whether quantitative, qualitative or mixed. However, few studies performed any kind of systematic analysis to evaluate the impacts of these strategies and were more descriptive in nature. The goal of a large portion of the studies was to share knowledge as quickly as possible, but the lack of rigorous analyses is problematic in identifying which strategies are effective.

An important influence in the interpretation of a strategy or a specific intervention was that most studies were written by healthcare workers working directly in a given hospital during the outbreak. Participation by healthcare workers in the process of knowledge creation can be an invaluable tool, demonstrating what Alexander et al. (2020) have identified as “reflexivity on action”, and enabling the creation of a “collective space for health professionals to reflect on and improve their practices”. However, this process could also represent a scientific bias that can bring into question the neutrality of the scientific research process, especially as many articles, particularly those in Chinese, did not consider the methodological limitations. Similarly, as China’s research and medical communities are not independent from politics (Chen, 2020), political factors may have played a role in the choice of papers written and published, potentially neglecting those that found negative results. These two factors: politics and the predominance of healthcare workers, as opposed to professional researchers, as authors, may also have limited the scope of articles concerning resilience issues such as finance or power structures, which can be sensitive and politicised.

Inequalities were largely ignored in the selected studies. Many of the articles that examined hospital strategies to address healthcare worker health issues emphasised the physical and mental health of nurses, while often neglecting the issues faced by other healthcare providers, including doctors. One possible reason for this phenomenon is that doctors may have more difficulty discussing problems encountered in work and sharing mental health concerns with colleagues (Galbraith et al. 2020). Similarly, gender issues and differences were not discussed in the selected papers. Despite significant gender gaps existing in healthcare professions-men being overrepresented in senior healthcare roles, and underrepresented in nursing staff-this was not considered in the selected articles. This suggests that a “gender blindness”, the systemic failure to acknowledge gender differences in health, (Smith, 2019; Connor et al. 2020) may be present in the case of Chinese hospitals.

These papers also highlighted how some processes undertaken during the pandemic attempted to increase healthcare access in ways that could potentially lead to a positive transformation process (as mentioned in the resilience framework). For example, articles focusing on e-health and ‘internet hospital’ interventions (Ding et al. 2020) mentioned ways that the transition to telemedicine provoked by the SARS-CoV-2 pandemic could be used to make healthcare more approachable, affordable and improve availability to vulnerable groups across the country. Further research is needed to examine whether these resilience processes have led to improved access to healthcare in China’s hospitals following the pandemic.

### 4.2 Recommendations for healthcare practitioners and managers

There are a number of recommendations offered to healthcare practitioners within the articles (Table 4). Improving patient awareness of online services enabled patients to better respond to these public health emergencies, and reducing unnecessary round trips between home and hospital (Wu, Guo et al. 2020, Lu, Hao et al. 2020). Artificial intelligence and internet technologies can be used for online self-assessment systems, robots can be used in guiding patients and delivering medicines within the hospital, and QR codes can be used for collecting patient and visitor information (Chen et al., 2020). Studies also found that China’s advanced use of technology has a crucial role in many elements of a resilience framework, including training, knowledge management and transfer and information systems. However, it is important to note that these recommendations were not substantiated by evidence. For example, Yan, Zou and Mirchandani (2020) provide a descriptive examination of information system strategies used by China’s most reputable hospitals and offer recommendations without any demonstration of evidence to support the recommendations.

In terms of nursing management, clear role recognition is seen as an important prerequisite for better practice. Nurses in one article (Wu et al., 2020) criticised the ambiguity of the roles given to over half of transdisciplinary nurses, suggesting that “more detailed role classification, clearer role definitions and job descriptions, and appropriate suggestions for expanded responsibilities would be effective methods to alleviate role ambiguity and improve work efficiency.” Other articles suggested that role ambiguity can be remedied with fairly simple interventions, such as a PDCA cycle to improve standardised nursing management in an ICU ward (Liu et al., 2020).

With regards to training, in articles that analyzed the impact of training interventions, both more traditional and online training interventions were associated with positive effects on knowledge and behaviour of staff regarding safety procedures, when compared to results before the training. Online or MOOC-based training is an appealing alternative to in-person training when reducing the risk of infection is a relevant concern. In order to build hospital resilience, articles argue that staff training for outbreak and infectious diseases practices should continue in regions without ongoing outbreaks (Gao, Sanna et al., 2020), and should continue after the outbreak has subsided (Chen et al., 2020). The ability to provide timely training interventions in response to a healthcare emergency is a crucial element of resilience framework.

Infection control measures comprise a crucial element of hospital resilience and many recommendations were given, although not always supported by data. Li and Chan et al., 2020 recommended targeting certain infection control interventions in low-risk departments, as there may be higher risk to staff on other wards (such as the maternity unit) compared to the infectious disease unit, due to disparate security measures and PPE use. Many articles related to infection control and environmental contamination recommended using risk-averse strategies with multiple layers of redundancy to reduce the risk of nosocomial and healthcare worker infection. Xu et al., (2020) also recommended that medical institutions should implement ward reconstruction, so that non-specialised hospital buildings are able to meet the requirements of an infectious disease unit.

### 4.3 Recommendations for Researchers

The results of these studies demonstrate the degree to which China’s healthcare system responded and adapted to the outbreak through several innovative measures. While evidence of the effectiveness of certain interventions was not provided, the collection of studies from across hospitals in China, offers strategies that together, have likely contributed to the drop in nosocomial infections from a peak of 127 new healthcare worker infections on the 23 Jan 2020, to the first day with 0 new cases on 8 March 2020 (Zheng, Wang et al. 2020).

Because of the short time frame, the lack of academic diversity in the research areas, political concerns and publication bias, this scoping review highlights the need for more rigorous intervention research and evaluation, and the inclusion of multi-disciplinary teams involving social science researchers and data scientists (Ridde et al., 2019). Gilson et al. (2020) have called for a structured research agenda to inform health policy and system responses to COVID-19, which should include resilience research in China’s hospitals.

This scoping review was not intended to draw conclusions about the causality of any particular strategy, therefore a ‘realist review’ (Pawson et al. 2005) would be a useful way of determining middle-range theories specific to the resilience of China’s hospitals in the outbreak.

### 4.4 Future Directions of Study

Our research also revealed that there are relatively few articles that have used the concept of resilience in a Chinese medical context, indicating that China’s hospitals do not consider a resilience framework as part of their research. Despite increased use in academic and professional contexts, the popular concept of *health systems resilience* has not yet reached conceptual maturity (Turenne et al., 2019) and, according to a recent scoping review “empirical studies fundamentally differ in the way that resilience is understood in a healthcare context” (Biddle et al. 2020). In China, the multiple possible translations for the term ‘resilience’: most prominently, *tanxing (ᐧ*性) *renxing (ᐧ*性) and *fuyuanli (ᐧ*原力), three terms with subtly different connotations, demonstrate this lack of conceptual clarity. Further research needs to be undertaken to understand how the concept of resilience translates and is understood across cultures and academic contexts.

Another element that must be addressed is the trade-offs associated with the risk-averse strategy employed in Chinese hospitals. Some studies noted that hospitals chose to implement a highly risk-averse strategy and that this did not allow them to determine what the minimal effective level of PPE use was to maintain effective protection (Liu et al., 2020), which poses problems for knowledge transfer to regions or situations with more limited capacity or resources. As Jin et al. (2020) note: “…(L)ack of evidence means we are using a precautionary approach which often results in our applying all available controls all the time”. Future comparative work could clarify whether China’s successes could be replicated without such extreme levels of personal protection, or whether a highly risk-averse, ‘zero-tolerance’ policy for nosocomial infection is the optimal choice.

Financial constraints, which comprise a central aspect of health systems resilience, have also been understudied in the Chinese context. While comparative studies have examined macro-level decisions and cost-benefit trade-offs in COVID-19 policy between countries including China (Balmford, 2020), our study found a lack of research pertaining to financial constraints faced by Chinese hospitals and other relevant decisions at the hospital level.

### 4.5 Limitations

We were unable to perform a risk-of-bias test for this paper, therefore the issues resulting from political or other biases were difficult to determine. As few of the articles were written by non-Chinese citizens or were peer-reviewed by external reviewers, selection effects caused by censorship cannot be excluded.

We chose not to include grey literature in this review, but it is worth noting that media articles, social media content and government white papers may also provide relevant sources of information that may help better understand how Chinese hospitals have sustained resilience during the SARS-CoV-2 outbreak.

## 5 Conclusion

Our scoping review demonstrates that there is a wide range of studies concerning hospital resilience in the Chinese context, and that this literature helps us greatly to understand the strategies used by the hospitals in China during the SARS-CoV-2 outbreak. The literature, both in Chinese and English, can provide important lessons on reinforcements, organization of work, e-health, telemedicine and use of technology, healthcare worker well-being, emergency team and nursing management, training, communication and information, protection protocols, PPE and reorganisation of services.

While this review demonstrates that the evidence is generally insufficient to determine the effectiveness of specific strategies, some preliminary results on the effectiveness of training interventions, technology use and management interventions, such as checklists and the PDCA cycle management, are provided. Furthermore, the study illuminates some common characteristics that have characterised what has generally been viewed as an effective strategy against the SARS-CoV-19 outbreak (Balmford et al. 2020), including risk aversion and redundancy.

## Data Availability

All data is available in supplementary files attached to the article.

## CRediT author statement

**Jack Stennett:** Formal analysis; Investigation; Data Curation, Writing - Original Draft; **Renyou Hou:** Formal analysis; Investigation; Data Curation, Writing - Original Draft; **Lola Traverson:** Project Administration, Writing - Review & Editing; **Valéry Ridde:** Project Administration, Conceptualization, Supervision, Writing-Review and Editing, Funding Acquisition; **Kate Zinszer:** Writing-Review and Editing**; Fanny Chabrol:** Writing-Review and Editing.

## Acknowledgments

We would like to sincerely thank Emmanuel Bonnet (CEPED), Laurence Goury (CEPED), and Loïc Min-Yu (BULAC) for their support in this research process.

